# Genetic Correlates of Treatment-Resistant Depression: Insights from Polygenic Scores Across Cognitive, Temperamental, and Sleep Traits in the All of US cohort

**DOI:** 10.1101/2024.07.03.24309914

**Authors:** Bohan Xu, Katherine L. Forthman, Rayus Kuplicki, Jonathan Ahern, Robert Loughnan, Firas Naber, Wesley K. Thompson, Charles B. Nemeroff, Martin P. Paulus, Chun Chieh Fan

**Affiliations:** Population Neuroscience and Genetics Center, Laureate Institute for Brain Research, Tulsa, Oklahoma, USA; Laureate Institute for Brain Research, Tulsa, Oklahoma, USA; Center for Human Development, University of California, San Diego, La Jolla, California, USA; Division of Biostatistics and Bioinformatics, the Herbert Wertheim School of Public Health and Human Longevity Science, University of California, San Diego, La Jolla, California, USA; Department of Psychiatry and Behavioral Sciences, Dell Medical School, The University of Texas at Austin, Austin, Texas, USA; Department of Psychiatry, University of California, San Diego, La Jolla, California, USA; Department of Radiology, University of California, San Diego, La Jolla, California, USA

**Keywords:** Treatment-resistant depression, polygenic scores, genetic predisposition, All of Us Research Program, major depressive disorder

## Abstract

**Background:** Treatment-resistant depression (TRD) is a major challenge in mental health, affecting a significant number of patients and leading to considerable economic and social burdens. The etiological factors contributing to TRD are complex and not fully understood.

**Objective:** To investigate the genetic factors associated with TRD using polygenic scores (PGS) across various traits, and to explore their potential role in the etiology of TRD using large-scale genomic data from the All of Us Research Program (AoU).

**Methods:** Data from 292,663 participants in the AoU were analyzed using a case-cohort design. Treatment resistant depression (TRD), treatment responsive Major Depressive Disorder (trMDD), and all others who have no formal diagnosis of Major Depressive Disorder (non-MDD) were identified through diagnostic codes and prescription patterns. Polygenic scores (PGS) for 61 unique traits from seven domains were used and logistic regressions were conducted to assess associations between PGS and TRD. Finally, Cox proportional hazard models were used to explore the predictive value of PGS for progression rate from the diagnostic event of Major Depressive Disorder (MDD) to TRD.

**Results:** In the discovery set (104128 non-MDD, 16640 trMDD, and 4177 TRD), 44 of 61 selected PGS were found to be significantly associated with MDD, regardless of treatment responsiveness. Eleven of them were found to have stronger associations with TRD than with trMDD, encompassing PGS from domains in education, cognition, personality, sleep, and temperament. Genetic predisposition for insomnia and specific neuroticism traits were associated with increased TRD risk (OR range from 1.05 to 1.15), while higher education and intelligence scores were protective (ORs 0.88 and 0.91, respectively). These associations are consistent across two other independent sets within AoU (n = 104,388 and 63,330). Among 28,964 individuals tracked over time, 3,854 developed TRD within an average of 944 days (95% CI: 883 ∼ 992 days) after MDD diagnosis. All eleven previously identified and replicated PGS were found to be modulating the conversion rate from MDD to TRD. Thus, those having higher education PGS would experiencing slower conversion rates than those who have lower education PGS with hazard ratios in 0.79 (80^th^ versus 20^th^ percentile, 95% CI: 0.74 ∼ 0.85). Those who had higher insomnia PGS experience faster conversion rates than those who had lower insomnia PGS, with hazard ratios in 1.21 (80^th^ versus 20^th^ percentile, 95% CI: 1.13 ∼ 1.30).

**Conclusions:** Our results indicate that genetic predisposition related to neuroticism, cognitive function, and sleep patterns play a significant role in the development of TRD. These findings underscore the importance of considering genetic and psychosocial factors in managing and treating TRD. Future research should focus on integrating genetic data with clinical outcomes to enhance our understanding of pathways leading to treatment resistance.

**Key Points:** *Question:* What are the predisposing characteristics among individuals who develop treatment-resistant depression (TRD)?

*Findings:* Analysis of data from 292,663 participants in the All of Us Research Program revealed that polygenic scores (PGS) for traits including neuroticism, cognitive function, and sleep patterns were significantly associated with major depressive disorder (MDD) and, particularly, with TRD. Among the 61 traits studied, 11 showed stronger associations with TRD compared to treatment responsive MDD, including traits linked to higher education and intelligence which appeared protective, and neuroticism and insomnia which increased risk.

*Meaning:* The findings underscore the importance of considering predisposing factors when managing and treating TRD. They suggest potential intervening pathways through tailored approach with the identified predisposing characteristics, reducing the risk of progression to treatment resistance in depression. Personalized genetic information that measures the underlying predispositions could eventually enhance therapeutic strategies.

## Introduction

Treatment Resistant Depression (TRD) is generally operationally defined as a major depressive disorder (MDD) with poor response to two trials of different classes of antidepressants ^1^. It exerts an enormous burden on quality of life ^2^ and healthcare resource utilization ^3^. Out of 8.9 million treated MDD patients in the US, about 2.8 million are estimated to have TRD, which amounts to an overall cost of almost $44 billion in treating these patients ^4^. Compared to treatment responsive individuals, those with TRD are 20% ^5^ to 30% ^6^ more expensive to treat, account for as much as 70% ^7^ more emergency department visits, outpatient visits, and prescriptions ^8^, and are 40% more likely to be hospitalized ^9^. These individuals have greater lost productivity ^10^, higher rates of permanent disability ^11^, and higher levels of suicide attempts and completed suicide ^12^.

Despite the clinical significance of TRD, the etiological factors remain elusive. TRD has been associated with higher prevalence of psychiatric comorbidities, including anxiety disorders^10^, stress disorders ^13^, and substance use disorder ^14^. Greater occurrences of ADHD, eating disorders, psychotic features, bipolarity, insomnia, and neuroticism are also reported ^15,16^. Histories of childhood maltreatment were found to be associated with the development of TRD ^17,18^, particularly among those who have genetic predispositions to psychiatric disorders ^19,20^, In regards to medical comorbidities , indicated associations include diabetes, immune system disorders, cardiovascular disease, and physical pain ^15,16^. Social factors are also implicated, as many demographic variables have been shown to be associated with TRD status ^15,16^. While the abundant literature points out the complex clinical features of TRD, it is unclear if those observed co-occurrences are the causes of difficult-to-treat depression. Given that depression is a potential risk factor for cardiovascular disease and diabetes ^21–23^, it is possible that long-lasting depressive symptoms lead to inflammatory processes shared by physical comorbidities. Misclassification of MDD may also lead to TRD. Furthermore, associated social factors may result from chronic debilitating effects of depression symptoms.

Understanding the causal mechanisms of TRD is of great importance to derive mechanism-based treatment approaches, and recent studies have employed genetically derived variables, such as polygenic scores (PGS), as causal instruments. The utilization of these genetically derived variables helps mitigate confounding factors due to the randomization in gametes, thereby providing insights into shared biological processes or potential mediating directions ^24^. For instance, in UK cohorts, PGS for major depression, schizophrenia, bipolar disorder, subjective wellbeing, intelligence, and neuroticism have shown no significant associations with TRD status among participants with MDD ^25^. While PGS for psychosis showed an association with TRD status when subgrouped with clinical characteristics, these findings have not been replicated in other cohorts ^25,26^. Moreover, genome-wide association studies (GWAS) of TRD have neither identified significant loci nor replicated any implicated candidate genes ^27^. Thus, these findings underscore the complexity of TRD’s genetic underpinnings and suggest the need for further research to clarify relationships.

To address the critical gap in understanding the etiology of TRD, we utilized the All of Us Research Program (AoU), a cohort drawn from hospital systems across all 50 US states that includes electronic health records, whole genome sequencing, and health survey responses ^28^. For this analysis, we selected 61 PGS representing unique traits across seven domains, which were derived from summary statistics of GWAS on samples independent of AoU (see Methods). These PGS were applied to the whole genome sequencing and microarray data of participants to critically examine their association with TRD status. Further validation of our findings was pursued through replication studies using two independent and non-overlapping cohorts within AoU.

## Methods

### All of Us

The cohort consisted of participants in the v7 release of the AoU Research Program. The AoU dataset includes electronic health records (EHR), whole genome data, physical measurements, and health questionnaires. The AoU data have been described previously ^29^. There are 413,457 participants in the v7 release. Participants were excluded if: (1) they indicated their sex at birth as *other*, (2) they live in a U.S. territory, (3) their state value was missing, (4) they had no EHR data, or (5) they did not have any genomic data. The final sample consisted of 292,663 participants in total.

We then separated the included samples into three independent and mutually exclusive cohorts, (1) participants who have short-read whole genome sequencing (WGS) and are genetically similar to persons of European ancestry (WGS European set, n = 124,945), (2) participants who have WGS and are genetically diverse (WGS Diverse set, n = 104,388), and (3) participants who did not have WGS but have been genotyped with the Illumina Infinium Global Diversity Array (Microarray set, n = 63,330). We deliberately chose these grouping factors because they were predefined without us preforming the random selection, enabling external replications. Details on the demographics for the three cohorts can be found in the **Table 1**.

**Table 1.** Demographic characteristics table.

|  |  | WGS, European set |  |  | WGS, Diversity set |  |  | Microarray |  |  |
| --- | --- | --- | --- | --- | --- | --- | --- | --- | --- | --- |
|  |  | non-MDD | trMDD | TRD | non-MDD | trMDD | TRD | non-MDD | trMDD | TRD |
|  |  | (N=104128) | (N=16640) | (N=4177) | (N=93881) | (N=8578) | (N=1929) | (N=55801) | (N=6106) | (N=1423) |
| **Sex** |  |  |  |  |  |  |  |  |  |  |
|  | Female | 59921 (57.5%) | 11554 (69.4%) | 2975 (71.2%) | 55820 (59.5%) | 6386 (74.4%) | 1492 (77.3%) | 32633 (58.5%) | 4207 (68.9%) | 993 (69.8%) |
|  | Male | 44207 (42.5%) | 5086 (30.6%) | 1202 (28.8%) | 38061 (40.5%) | 2192 (25.6%) | 437 (22.7%) | 23168 (41.5%) | 1899 (31.1%) | 430 (30.2%) |
| **Birth Year** |  |  |  |  |  |  |  |  |  |  |
|  | Median (Min, Max) | 1961<br>(1905, 2004) | 1960<br>(1918, 2003) | 1962<br>(1926, 2003) | 1972<br>(1915, 2004) | 1966<br>(1923, 2003) | 1966<br>(1930, 2002) | 1966<br>(1901, 2004) | 1962<br>(1921, 2004) | 1965<br>(1924, 2001) |
| **Self-report Race** |  |  |  |  |  |  |  |  |  |  |
|  | Asian | 30 (0.0%) | 6 (0.0%) | 0 (0%) | 6975 (7.4%) | 338 (3.9%) | 63 (3.3%) | 2096 (3.8%) | 64 (1.0%) | 14 (1.0%) |
|  | Black or African American | 89 (0.1%) | 17 (0.1%) | 3 (0.1%) | 43226 (46.0%) | 3852 (44.9%) | 961 (49.8%) | 10599 (19.0%) | 1058 (17.3%) | 289 (20.3%) |
|  | Middle Eastern or North African | 451 (0.4%) | 45 (0.3%) | 10 (0.2%) | 724 (0.8%) | 65 (0.8%) | 20 (1.0%) | 395 (0.7%) | 31 (0.5%) | 7 (0.5%) |
|  | Native Hawaiian or Other Pacific Islander | 22 (0.0%) | 1 (0.0%) | 0 (0%) | 198 (0.2%) | 18 (0.2%) | 3 (0.2%) | 88 (0.2%) | 5 (0.1%) | 0 (0%) |
|  | Others | 2188 (2.1%) | 333 (2.0%) | 82 (2.0%) | 3320 (3.5%) | 361 (4.2%) | 87 (4.5%) | 1593 (2.9%) | 185 (3.0%) | 54 (3.8%) |
|  | White | 98730 (94.8%) | 15820 (95.1%) | 3975 (95.2%) | 3690 (3.9%) | 389 (4.5%) | 81 (4.2%) | 30781 (55.2%) | 3816 (62.5%) | 877 (61.6%) |
|  | Missing | 2618 (2.5%) | 418 (2.5%) | 107 (2.6%) | 35748 (38.1%) | 3555 (41.4%) | 714 (37.0%) | 10249 (18.4%) | 947 (15.5%) | 182 (12.8%) |
| **Self-report Ethnicity** |  |  |  |  |  |  |  |  |  |  |
|  | Hispanic or Latino | 1598 (1.5%) | 181 (1.1%) | 41 (1.0%) | 37846 (40.3%) | 3835 (44.7%) | 751 (38.9%) | 10584 (19.0%) | 945 (15.5%) | 179 (12.6%) |
|  | Not Hispanic or Latino | 99913 (96.0%) | 15981 (96.0%) | 4013 (96.1%) | 53252 (56.7%) | 4487 (52.3%) | 1115 (57.8%) | 43723 (78.4%) | 4982 (81.6%) | 1187 (83.4%) |
|  | Others | 884 (0.8%) | 161 (1.0%) | 37 (0.9%) | 1108 (1.2%) | 122 (1.4%) | 27 (1.4%) | 564 (1.0%) | 61 (1.0%) | 25 (1.8%) |
|  | Missing | 1733 (1.7%) | 317 (1.9%) | 86 (2.1%) | 1675 (1.8%) | 134 (1.6%) | 36 (1.9%) | 930 (1.7%) | 118 (1.9%) | 32 (2.2%) |

\* non-MDD, individuals who did not have formal diagnosis of Major Depressive Disorder in record; trMDD, treatment responsive
Major Depressive Disorder; TRD, treatment resistant depression.

### Determining Status

We operationalized TRD based on a participant’s engagement with three or more distinct antidepressant drugs within a single drug trial period ^30^. Participants might experience multiple distinct drug trial periods throughout their medical history, thus, each of these periods were evaluated independently based on the criteria set forth for the index drug and subsequent treatment within a one-year frame. This definition was consistent with the approach used in the UK ^25,27^, Sweden ^31^, and Taiwan ^32^. This definition took into account the dynamic nature of antidepressant treatment strategies over time, allowing us to systematically assess treatment patterns and the incidence of TRD across the participant cohort.

The characterizations of treatment-responsive MDD (trMDD), TRD, and MDD negative (non-MDD) were determined by diagnostic codes recorded in the EHR and the medication record. The average length of EHR was 11.89 years (SD = 8.98 years, Median = 8.95 years, Range = [0.003, 93.07] years). MDD status was determined based on presence of a diagnostic entry from a list of diagnostic codes in the International Classification of Diseases (ICD v9 and v10, see **Supplementary eTable 1**). Subsequently, we identified the drug trial period for everyone with MDD, defined as a continuous treatment period that begins with the first prescription or refill entry and ends with a gap of more than 6 months without record of a subsequent prescription. We standardized the prescription records to focus the analysis on the occurrence and continuity of antidepressant use rather than the specifics of each prescription, such as dosage or formulation.

### Polygenic Scores

We selected 61 summary statistics from seven domains to generate the corresponding PGS: (a) Education and cognition (2 PGS)^33^, (b) Metabolic, somatic complaints, and inflammation traits (17 PGS)^34–38^, (c) Personality (19 PGS)^39–43^, (d) Psychiatric disorders (9 PGS)^40,43–49^, (e) Sleep patterns (2 PGS)^43^, (f) Substance use (6 PGS)^43,50^, (g) Temperament ^40^ (6 PGS). Details of each selected PGS, including publication records, can be found in the **Supplementary eTable 2**. We selected these PGS based on previous studies reporting that their corresponding observed traits were associated with TRD^15,16^.

We used PRS-CS, a continuous prior based shrinkage method ^51^, to generate the posterior weights given the GWAS summary statistics. To avoid directly using AoU for calibrating weights, we utilized a smaller, locally available genomic dataset, the Tulsa 1000 study (T1000) ^52^. T1000 is a longitudinal study including 1000 individuals with mood/anxiety, substance use, or eating disorders, and healthy controls. The genotyping was done with the Illumina Infinium Global Screening Array-24 (v.2.0) and imputation was performed via the Michigan Imputation Sever Pipeline (Minimac4, version 1.2.4), using the HRC reference panel ^53^. PRS-CS estimates global and local scaling parameters by simultaneously evaluating the association strengths of a group of SNPs in a linkage disequilibrium block and their correlation patterns in the reference genotype data (in this case, the T1000). Those estimated parameters were later used as shrinkage factors to determine the posterior effect sizes, which we used for calculating PGS in AoU.

To calculate the PGS of each trait in AoU accordingly, we applied the posterior effect sizes to the Allele Count/Allele Frequency (ACAF)-thresholded short-read sequencing data provided by AoU. This data was filtered based on a preset ACAF threshold, which required that either the population-specific allele frequency (AF) exceeded 1% or the population-specific allele count (AC) was greater than 100 in any of the ancestry subpopulations. We then excluded sites based on four criteria: (a) excess heterozygosity, (b) overall AF of 0.5% or less, (c) multi-nucleic alleles, or (d) a call rate under 99%. We ended with 10,222,713 SNPs after applying these filters. We applied the calculated posterior effect sizes to the WGS of AoU, generating the PGS of each trait accordingly.

For the Microarray set, we calculated the PGS based on all available SNPs from the Illumina Infinium Global Diversity Array. The total number of available SNPs was 1,739,268. To serve as a replicating analysis, we directly apply the posterior effect sizes to the intersecting SNP sets to generate the PGS of each trait.

### Statistical Methods

First, we used logistic regression to determine the associations between each PGS and each binary diagnostic status, amounting to three outcome comparisons conducted for each PGS. Diagnostic comparisons included trMDD versus non-MDD, TRD versus non-MDD, and TRD versus trMDD. In each regression, we include 16 genetic principal components, biological sex, and age as the covariates. For discovery, significance threshold was determined at 0.0001, the Bonferroni correction for two-tail tests on 61 PGS with three different outcome contrasts. Effect measures (odds ratios, OR) and the corresponding 95% confidence intervals are also reported. For replication, we examined if the point estimates of the all the associations were consistent across pre-selected independent cohorts.

To determine whether PGS were associated with progression from MDD to TRD, time-to-event analyses were performed on the PGS that significantly predicted diagnostic category. We selected a subset of patients with major depressive disorders according to the three previously described cohorts, and included only those who have more than two diagnostic time points on record (n = 19124, 9840, 6967 for WGS European set, WGS Diverse set, and Microarray set, respectively). We applied a Cox proportional hazard model to estimate the progression rate given the PGS while controlling for age at MDD diagnosis, sex, and 16 principal components of genetic ancestry. Hazard ratios and their corresponding 95% confidence intervals were estimated. The proportional hazard assumptions were examined via both a graphic method and a Schoenfeld test.

## Results

### PGS-associated risk of developing TRD

**Figure 1a** summarizes the associations between each PGS and disease status. In the WGS European set, 44 of our selected 61 PGS show significant associations with the likelihood of being trMDD versus non-MDD (all P_bonferroni_ < 0.05). The validated list includes 2 PGS from Education and Cognition, 10 PGS from Metabolic, somatic complaints, and inflammations, 17 PGS from Personality, 5 PGS from Psychiatric disorders, 1 PGS from Sleep, 3 PGS from Substance use, and 6 PGS from Temperament (**Supplementary eTable 3**). On average, the point estimates of the OR have larger magnitude in TRD-vs-non-MDD than in trMDD-vs-non-MDD. However, TRD-vs-non-MDD have larger confidence intervals than trMDD-vs-non-MDD due to the reduced number of defined cases. To see if the overall association patterns hold in the replication set while reduce the impact of limited sample sizes, we performed meta-analytic correlations to compare the effect size estimates across sets. We found that the association patterns are highly consistent. Meta-analytic correlations between the WGS Diverse set and European set are 0.89 and 0.83 for trMDD-vs-non-MDD and TRD-vs-non-MDD, respectively (**Supplementary eFigure 1 and Supplementary eFigure 2**).

**Figure 1.**
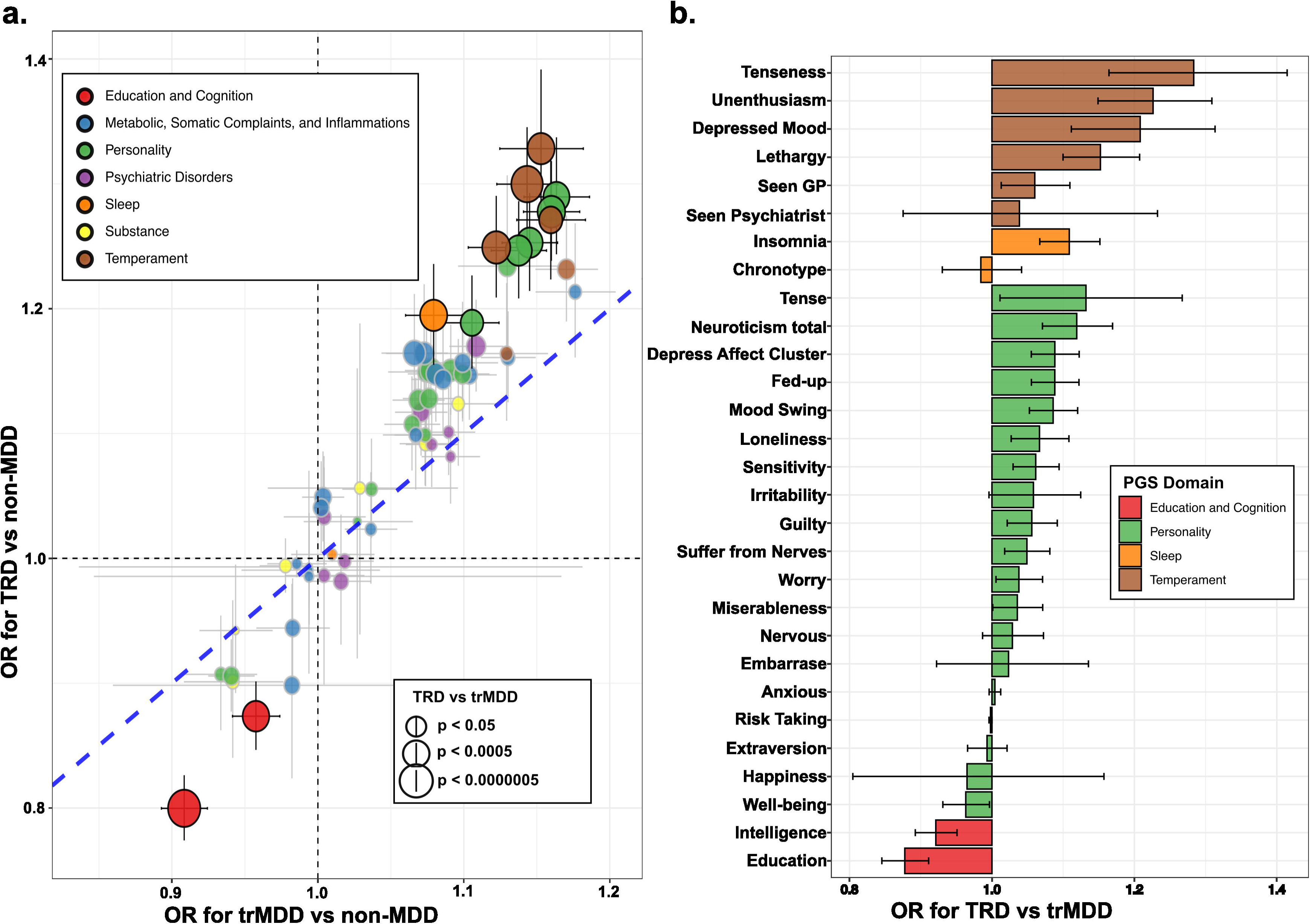
Associations between PGS and diagnostic status. (a) The scatter plots show the overall effect sizes of every included PGS, with the corresponding 95% confidence intervals. X-axis represents the odds ratios (OR) in comparing trMDD to non-MDD groups, for every increase one standard deviation of PGS. Y-axis represents the OR in comparing TRD to non-MDD groups, for every increase one standard deviation of PGS. PGS that show significant odds ratios in being TRD-vs-trMDD, after Bonferroni correction, are highlighted in solid colors. (b) The distribution of the odds ratios in the domains where individual PGS show significant stronger associations with TRD status than with trMDD.

Despite a high degree of similarity in the association patterns, 11 PGS showed stronger associations with TRD than with trMDD in both European and Diversity cohorts (P_bonferroni_ < 0.05, Figure 1a and 1b). Those 11 PGS belong to four different domains: Education/Cognition, Sleep, Personality, and Temperament. Some of the significant PGS in Temperament stand for trans-diagnostic psychiatric symptoms, such as lethargy, depressed mood, and tenseness in the past two weeks. However, none of the PGS for psychiatric disorders, including PGS for Major Depressive Disorder, were shown to be significantly associated with TRD status among patients with MDD, neither did PGS for Substance Use and PGS for Metabolic, Somatic Complaints, and Inflammation. Figure 1b shows the distribution of the OR and the corresponding 95% confidence intervals for TRD vs trMDD for every one standard deviation (SD) difference in the PGS in the combined European and Diversity cohorts. Genetic propensity for depressive affect, including tenseness, unenthusiasm, depressed mood, and lethargy, increase the likelihood of being TRD by 28% (95%CI: 1.16 ∼ 1.41), 23% (95%CI: 1/14 ∼ 1.30), 21% (95%CI: 1.11 ∼ 1.31), and 15% (95%CI: 1.10 ∼ 1.209), respectively. Insomnia increases TRD risk by 11% (95%CI: 1.06 ∼ 1.15). Neuroticism and its item-level sub-scores (depressive affect cluster score, fed-up, mood swing, and loneliness) all increase the likelihood of being TRD, with neuroticism indicating a 11% increase in risk and ORs of the sub-scores ranging from 1.06 to 1.08. PGS predicting higher educational attainment and intelligence are associated with lower prevalence of TRD, with OR in 0.88 (95%CI: 0.84 ∼ 0.91) and 0.91 (95%CI: 0.89 ∼ 0.95), respectively. Estimates derived independently for the European and Diversity cohorts were consistent (see **Supplementary eFigure 3**).

These results are replicated in the Microarray set, despite fewer SNPs and fewer individuals compared to the WGS dataset (**Supplementary eFigure 4**). The meta-analytic correlations for TRD-vs-trMDD for all 61 PGS is 0.78. PGS of intelligence, education attainment, and insomnia remain significantly associated with TRD status despite greatly reduced sample sizes and input SNPs (P_bonferroni_ < 0.05).

### Predicting the progression from MDD to TRD

Among 28,964 individuals from the WGS set who had at least two time points, 3854 converged to TRD, on average, within 944 days after receiving MDD diagnosis (95% CI: 883 ∼ 992 days). We first examined whether those 11 PGS identified in the previous step could differentiate the time-to-TRD onset among patients with MDD. As showcased in Figure 2a, when individuals are stratified by the PGS of educational attainment (PGS_edu_), higher level of PGS_edu_ is associated with a slower progression rate to TRD than those who have lower level of PGS_edu_ (Kaplan-Meier curves for each PGS strata and corresponding 95% CI). We formally tested the time-modulating effects of those PGS using Cox proportional hazard models. Figure 2b shows the hazard ratios (HR) estimated from the time-to-TRD-onset analyses in the longitudinal WGS sets. After controlling for age-at-MDD diagnosis, sex, and first 16 genetic PCs, all 11 PGS show significant associations with time-to-TRD-onset. After Bonferroni correction, all except two sub-items of neuroticism retain significance. For instance, higher PGS_edu_ (80 percentile) is associated with slower progression rate compared to lower PGS_edu_ (20 percentile) (HR = 0.79 | 95% CI: 0.74 ∼ 0.85). Higher PGS for insomnia (80 percentile) is associated with a faster progression rate compared to lower PGS for insomnia (20 percentile) (HR = 1.21 | 95% CI: 1.13 ∼ 1.30). These results suggest that genetic propensity of those traits is not only associated with whether or not the individuals would have TRD but also with when to expect the TRD would occur. We repeated the analyses on the Microarray subset (n = 6967) and found virtually identical results (**Supplementary eFigure 5**).

**Figure 2.**
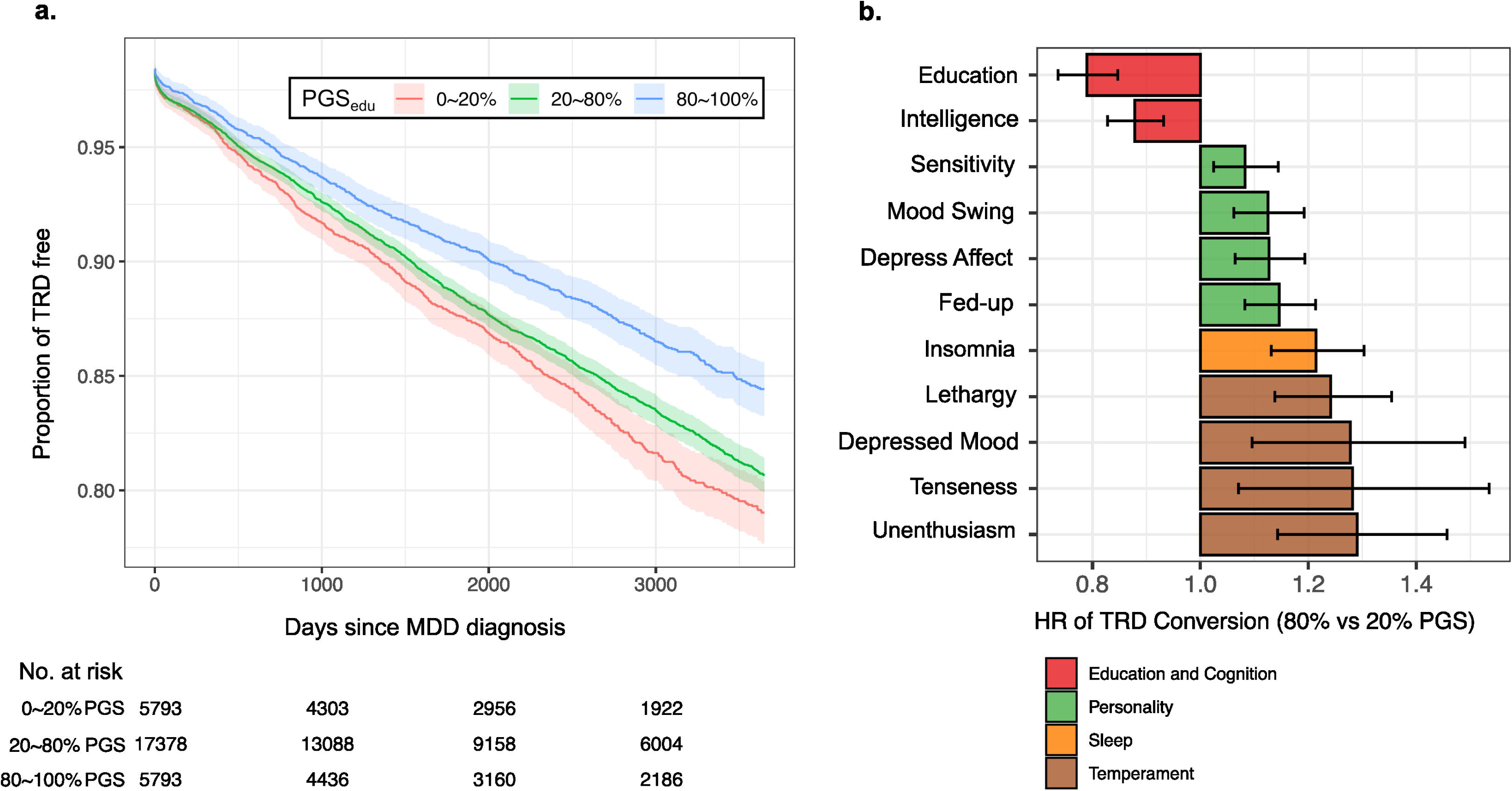
Progression rates of TRD from the first MDD diagnosis. (a) Kaplan-Meier curve of progression from MDD to TRD, stratified by the PGS of education attainment (PGS_edu_). Individuals were grouped by their PGS, as 0 to 20 percentile, 20 to 80 percentile, and 80 to 100 percentile. (b). Hazard ratios and the corresponding 95% confidence interval given PGS strata, estimated by Cox regression model, controlling for sex, age at diagnosis, and first 16 genetic PCs.

## Discussion

This investigation aimed to elucidate the etiology of TRD by leveraging the extensive AoU cohort and analyzing 61 polygenic scores from unique traits across seven domains. These PGS were derived from summary statistics of GWAS on samples independent of AoU and applied to the genotype data from three naturally occurring, mutually exclusive, independent cohorts in AoU. Our findings showed that increased likelihood (OR) of TRD is genetically correlated with traits such as depressive affect, neuroticism, and insomnia. Moreover, these genetic predictors were associated with progression from MDD to TRD (HR). These findings support that these traits are etiological factors for TRD ^15,16^. Conversely, traits related to intelligence and education showed opposite effects in both OR and HR, indicating a protective effect against TRD. The replication of these analyses in two independent AoU data sets confirmed the consistency of these associations, underscoring the robustness of our results. This study not only highlights the genetic predispositions linked to TRD—it also demonstrates the complex interplay of traits affecting the development of TRD, emphasizing the need for tailored intervention strategies to manage TRD effectively.

Our study found that genetic propensity for psychiatric conditions had no discriminative power to differentiate between individuals resistant to treatment and those responsive to treatment for MDD. This finding echo results from a similar study conducted with UK samples, where—despite a substantial sample size—no significant associations were found between TRD and PGS of psychiatric traits ^25^. This suggests that the higher levels of comorbidities observed in conditions like anxiety, stress, and psychotic disorders among treatment-resistant patients are not simply due to misclassification or shared etiological factors across different diagnoses. Rather, these comorbidities likely reflect the greater severity of depressive symptoms in patients who develop treatment resistance, underscoring the complexity of diagnosing and treating these severe forms of depression.

We found associations between metrics of neuroticism and TRD. Neuroticism is a personality trait that reflects a tendency to experience negative emotions ^54^, such as anxiety, depression, anger, and fear, in response to stressors or challenges ^55^. Neuroticism is approximately 40% ^56^ heritable, and high levels of neuroticism have been associated with many poor mental health outcomes ^57^. While neuroticism has been reported to be associated with TRD, the associations at the phenotypic level are not universally observed and mostly attributed to the diagnosis of MDD ^15,16^. Our results highlight the significant role neuroticism plays in both the onset and treatment resistance of MDD. Notably, strong associations with TRD in our result support that specific subtypes of neuroticism, such as the depressive affect cluster, have been linked to more severe progression of MDD ^25,58–65^. This underscores the shared genetic components between the depressive tendencies as a personality trait and the severity of the depressive symptoms as a treatment outcome. The genetic overlap between neuroticism and other psychiatric conditions^41,64–66^ highlights the need for a transdiagnostic approach to understanding and treating TRD.

In addition to the genetic pleiotropy on the depressed affects and depressive symptoms, it is also possible that high neuroticism scores correlate with poorer treatment response and greater treatment resistance are mediated by cognitive impairments such as reduced cognitive flexibility. Neuroticism, characterized by a higher sensitivity to negative stimuli ^67^, a propensity for rumination ^68^, and a reduced ability to regulate emotions ^69^, is deeply intertwined with cognitive and social processes that exacerbate depressive disorders. Neuroticism could be a key factor in not only the manifestation of MDD but also in its resistance to conventional treatments, making it a critical target for intervention strategies.

Furthermore, we also found PGS for educational attainment exhibits the strongest association with TRD status compared to other traits analyzed, with a weaker but consistent association in PGS for cognitive function. Such findings align with previous studies ^15,16,70,71^ that have noted phenotypic associations between higher education levels and reduced TRD risk, suggesting that these associations may have a shared genetic foundation potentially influenced by limited options in decision-making processes. This could explain why individuals with lower educational levels and reduced cognitive functions might experience poorer adherence to medication regimens ^72,73^, potentially leading to more frequent changes in antidepressant treatments. The evidence highlights the importance of considering cognitive factors in the management and understanding of TRD, emphasizing the need for further investigation into how these genetic predispositions impact treatment outcomes and resistance.

Our findings show that insomnia, rather than sleep chronotype, is significantly associated with the risk of TRD. Sleep disturbances are deeply intertwined with the depressive symptoms and have been a key feature among patients with TRD ^31,74,75^. While there are evidence supporting the notion that treating insomnia could alleviate depressive symptoms ^75^, pharmacological intervention with benzodiazepine is deemed to be inadequate or even harmful to patients with TRD ^74,76^. The association between PGS for insomnia and TRD in our results may be driven by a shared etiological mechanism between those two. This might explain why direct pharmacological boost to the sleep itself did not benefit patients whereas psychotropic agent, such as ketamine, can ameliorate both the depressive symptoms and sleep disturbance among patients with TRD simultaneously ^77^. Nonetheless, given that genetic propensities toward insomnia is associated with accelerated progression rate from MDD to TRD, we should still consider incorporating comprehensive insomnia management into the therapeutic strategies for MDD. Addressing insomnia may not only target a key symptom of depression but also enhances overall treatment efficacy, preventing the emergence of treatment resistance.

Our study has limitations.

The TRD status is determined by algorithm using EHR for drug events. We did not have dimensional measures on the depressive symptoms, treatment information other than drug prescriptions, and the actual adherence among patients. Those factors might lead to misclassifications between TRD and trMDD, blurring the diagnostic boundaries and reducing the statistical power to detect associations in TRD-vs-trMDD. Our estimates on the prevalence of TRD are consistent with the expected rates in national surveys ^4^ and we still found eleven PGS significantly associated with TRD status, indicating limited impact of the potential misclassifications. The statistical power of the PGS vary because of the differences in the sample size, training populations, and the genetic architecture of the phenotypic traits ^24,78^. The comparisons across PGS are not solely driven by the relationships between the traits of the PGS and TRD status. However, the PGS of psychiatric disorders that have higher heritability, and larger GWAS sample size do not show significant associations in our results, suggesting our results are not completely driven by the differences in the original GWAS. Finally, PGS captures genetic propensity, which does not fully determine the actual exposure histories to the environment, such as early life adversity that is known to have a substantial, and often larger, contribution to mental health outcomes ^17,18,20^. This may explain why the PGS for CRP did not differentiate the treatment responsiveness in our result despite evidence on the role of stress in the etiology of depressive symptoms ^20,79^. Since we did not actually measure the stress or inflammatory markers, it is possible that the actual experience of the inflammatory inducing events is more important than the physical predispositions toward inflammation in the genesis of TRD. Our findings should be interpreted as, after mitigating the potential bias and confounds based on the genetic instruments, the cognitive functions, neuroticism, general affect, and sleep disturbance play a more salient role in the treatment efficacy than the other conditions we included, implicating a clinical path forward to obtaining treatment responses among patients with MDD.

In conclusion, this comprehensive investigation into the etiology of TRD via the analysis of polygenic scores across diverse traits has brought to light several genetic factors that may influence the development and management of TRD. Our findings indicate strong genetic associations with traits such as tenseness, unenthusiasm, depressed mood, and lethargy—suggesting their potential as determinants for predicting TRD risk. Moreover, the negative associations observed with traits related to higher educational attainment and general intelligence point to potential protective factors, underscoring the complexity of TRD’s genetic landscape. The consistency of these findings across independent data sets enhances the robustness of our conclusions. Moreover, the correlation between high levels of neuroticism and TRD suggests that personality traits significantly contribute to the severity of depression. Importantly, the identification of insomnia as a treatable risk factor offers a viable pathway for clinical intervention. These insights not only advance our understanding of the genetic underpinnings of TRD, but also highlight critical areas for future research and potential therapeutic targets, ultimately aiming to improve treatment strategies and outcomes for those suffering from this challenging condition.

## Data Availability

All data produced in the present study are available upon reasonable request to the authors.

## Acknowledgement

This work was partly funded by The William K. Warren Foundation, the National Institute of General Medical Sciences Center (Grant 2 P20GM121312, MPP, RK, KLF), the National Institute on Drug Abuse (U01DA050989, MPP), and the National Institute for Mental Health (R01MH122688, R01MH128959, CCF). Dr. Nemeroff is supported by the National Institutes of Health, the National Institute of Mental Health, and the National Institute of Alcohol Abuse and Alcoholism.

## Notes

### Competing Interest Statement

Dr. Paulus advises Spring Care, Inc., receives royalties from an article on methamphetamine in
UpToDate, and has a compensated consulting agreement with Boehringer Ingelheim
International GmbH.
Dr. Nemeroff is a consultant for ANeuroTech (division Anima BV), Signant Health, Janssen
Research and Development, BioXcel Therapeutics, Silo Pharma, Engrail Therapeutics, Clexio Biosciences LTD, EmbarkNeuro, Galen Mental Health LLC, Goodcap Pharmaceuticals, ITI Inc,
LUCY Scientific Discovery, Relmada Therapeutics, Sage Therapeutics, Senseye Inc, Precisement
Health, Autobahn Therapeutics Inc, EMA Wellness, Skyland Trails, and the Brain & Behavior
Research Foundation. Charles Nemeroff owns the following patents: Method and devices for
transdermal delivery of lithium (US 6,375,990B1), Method of assessing antidepressant drug
therapy via transport inhibition of monoamine neurotransmitters by ex vivo assay (US
7,148,027B2), Compounds, Compositions, Methods of Synthesis, and Methods of Treatment
(CRF Receptor Binding Ligand) (US 8,551, 996 B2). Charles Nemeroff owns stock in Seattle
Genetics, Corcept Therapeutics Company, EMA Wellness, Precisement Health, Relmada
Therapeutics, Signant Health, Galen Mental Health LLC, and Senseye Inc.

### Author Declarations

The human data are available via The All of Us Research Program (https://allofus.nih.gov/)

