## Supplemental Figures and Tables for "Genetic Correlates of Treatment-Resistant Depression: Insights from Polygenic Scores Across Cognitive, Temperamental, and Sleep Traits in the All of US cohort"

**Supplementary Materials**

**eFigure 1. Replications of the trMDD vs non-MDD comparisons in the diverse set of WGS samples.**

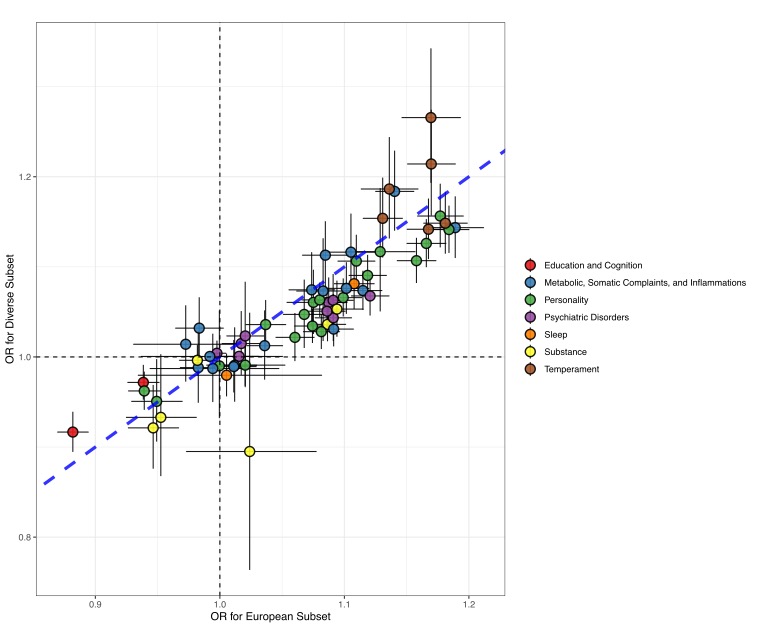

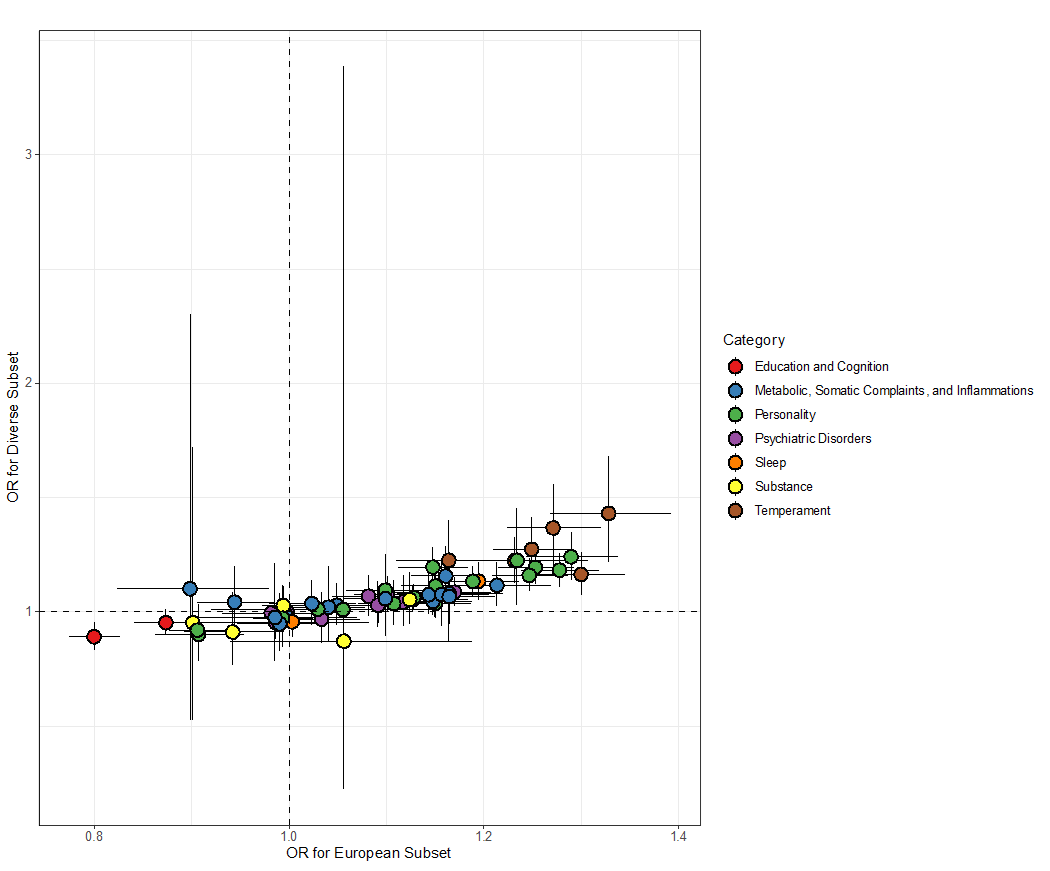
**eFigure 2. Replications of TRD vs non-MDD comparisons in the diverse set of WGS samples.**

**eFigure 3. Replications of TRD vs trMDD comparisons in the diverse set of WGS samples.**

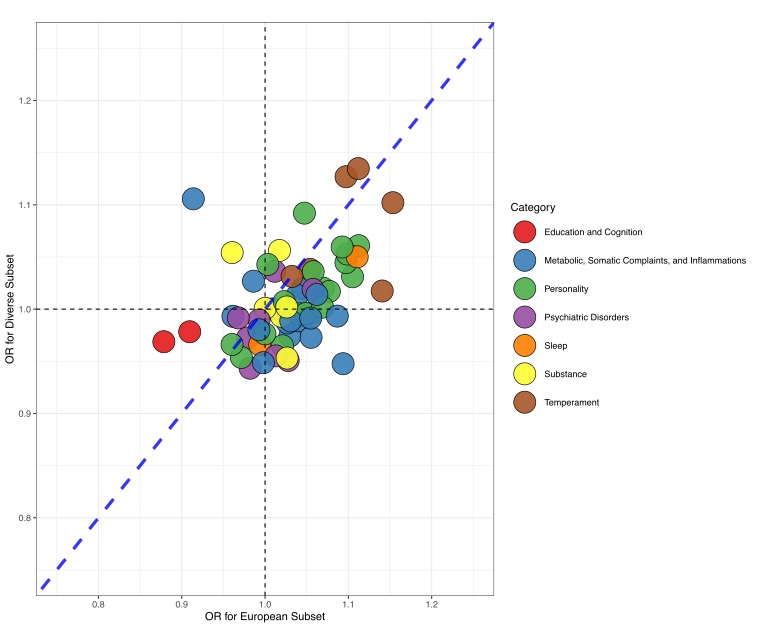

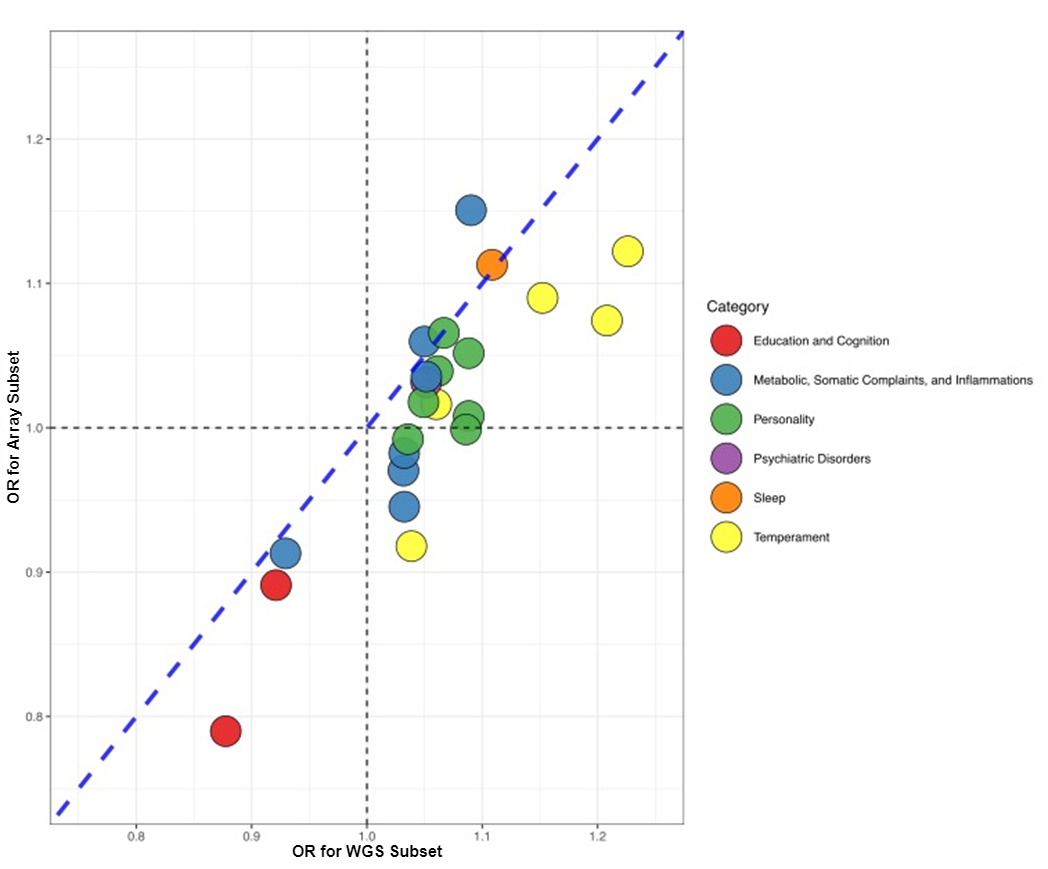
**eFigure 4. Replications of TRD vs trMDD comparisons in the microarray samples.**

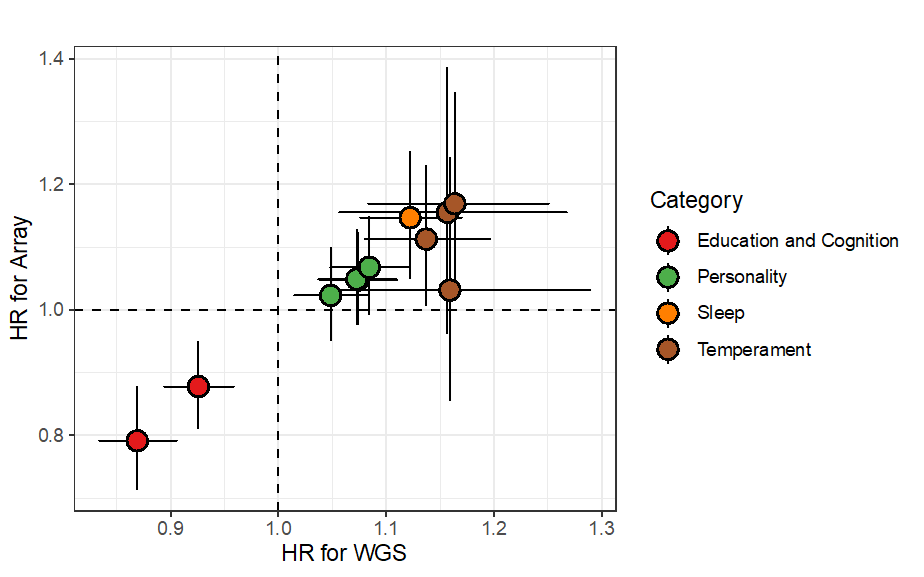
**eFigure 5. Comparison of hazard ratios for transition from MDD to TRD.**

**eTable 1. Selection criteria for MDD status using ICD-9 and ICD-10 codes.**

| **Inclusion Criteria** | |
| --- | --- |
|  | *Depression* |
| ICD-9 | 296.2, 296.3, 311 |
| ICD-10 | F32.0, F32.1, F32.2, F32.3, F32.4, F32.5, F32.8, F32.9, F32.89, F33.0, F33.1, F33.2, F33.3, F33.4, F33.8, F33.9, F33.40, F33.41, F33.42 |
| **Exclusion Criteria** | |
|  | *Psychosis* |
| ICD-9 | 291.8, 291.9, 295.0, 295.2, 295.3, 295.10, 295.11, 295.12, 295.14, 298.0, 298.1, 298.4, 298.8, 298.9, 299.9, 797, V11.0 |
| ICD-10 | F20.81, F20.89, F20.9, F25.0, F25.1, F25.9, F29, F53 |
|  | *Bipolar Disorder* |
| ICD-9 | 296.5, 296.6, 296.7, 296.8 |
| ICD-10 | F30.2, F30.8, F30.9, F30.10, F31.0, F31.2, F31.3, F31.30, F31.31, F31.32, F31.4, F31.5, F31.6, F31.7, F31.10, F31.11, F31.12, F31.13, F25.0 |

**eTable 2. An in-depth summary of 61 selected PGS.**

| **Category** | **PMID** | **Year** | **uniqTrait** | **Population** | **N** | **SNP.h2** | **Filename** | **Download link** |
| --- | --- | --- | --- | --- | --- | --- | --- | --- |
| Education and Cognition | 30038396 | 2018 | Educational attainment | UKB2 (EUR meta) | 766345 | 0.1055 | GWASA_ID_4066-Educational_Attainment | https://www.dropbox.com/s/ho58e9jmytmpaf8/GWAS_EA_excl23andMe.txt?dl=1 |
|  | 30038396 | 2018 | Intelligence | UKB2 (EUR meta) | 257828 | 0.1922 | GWASA_ID_4067-Cognitive_Performance | https://www.dropbox.com/s/ibjoh0g5e3sdd8t/GWAS_CP_all.txt?dl=1 |
| Metabolic, Somatic Complaints, and Inflammations | 31427789 | 2019 | Back pain | UKB2 (EUR) | 385698 | 0.0349 | GWASA_ID_3572-Back_Pain | https://atlas.ctglab.nl/ukb2_sumstats/6159_4_logistic.EUR.sumstats.MACfilt.txt.gz |
|  | 30239722 | 2018 | Body Mass Index | UKB2 (EUR meta) | 806834 | 0.1755 | GWASA_ID_4074-Body_Mass_Index | https://zenodo.org/record/1251813/files/bmi.giant-ukbb.meta-analysis.combined.23May2018.txt.gz?download=1 |
|  | 25010111 | 2014 | Cortisol | EUR | 12597 | NA | CORNET-2014Cortisol | https://datashare.is.ed.ac.uk/bitstream/handle/10283/2787/gwama_1_fixed.out?sequence=1&isAllowed=y |
|  | 6218410 | 2018 | CRP | EUR | 204402 | NA | HapMap_GWAS_CRP_AJHG | https://wetransfer.com/downloads/5f1a0bdfb4164c6432d85d6e650e264320200512145400/e277e1bd961090795966ff01bf2ed7d720200512145412/7eccdf |
|  | 31427789 | 2019 | Hayfever, allergic rhinitis or eczema | UKB2 (EUR) | 385822 | 0.0611 | GWASA_ID_3553-Hayfever_or_Allergic_Rhinitis_or_Eczema | https://atlas.ctglab.nl/ukb2_sumstats/6152_9_logistic.EUR.sumstats.MACfilt.txt.gz |
|  | 31427789 | 2019 | Headache | UKB2 (EUR) | 385698 | 0.0402 | GWASA_ID_3570-Headache | https://atlas.ctglab.nl/ukb2_sumstats/6159_1_logistic.EUR.sumstats.MACfilt.txt.gz |
|  | 31427789 | 2019 | Height | UKB2 (EUR) | 385748 | 0.3097 | GWASA_ID_3187-Height | https://atlas.ctglab.nl/ukb2_sumstats/f.50.0.0_res.EUR.sumstats.MACfilt.txt.gz |
|  | 24097068 | 2013 | High-density lipoprotein cholesterol | EUR | 188577 | 0.1598 | GWASA_ID_71-High_density_lipoprotein_cholesterol | http://csg.sph.umich.edu/abecasis/public/lipids2013/jointGwasMc_HDL.txt.gz |
|  | 31427789 | 2019 | Hip pain | UKB2 (EUR) | 385698 | 0.0223 | GWASA_ID_3574-Hip_Pain | https://atlas.ctglab.nl/ukb2_sumstats/6159_6_logistic.EUR.sumstats.MACfilt.txt.gz |
|  | 31427789 | 2019 | Knee pain | UKB2 (EUR) | 385698 | 0.0373 | GWASA_ID_3575-Knee_Pain | https://atlas.ctglab.nl/ukb2_sumstats/6159_7_logistic.EUR.sumstats.MACfilt.txt.gz |
|  | 24097068 | 2013 | Low-density lipoprotein cholesterol | EUR | 188577 | 0.1363 | GWASA_ID_70-Low_density_lipoprotein_cholesterol | http://csg.sph.umich.edu/abecasis/public/lipids2013/jointGwasMc_LDL.txt.gz |
|  | 31427789 | 2019 | Neck or shoulder pain | UKB2 (EUR) | 385698 | 0.0317 | GWASA_ID_3571-Neck_or_Shoulder_Pain | https://atlas.ctglab.nl/ukb2_sumstats/6159_3_logistic.EUR.sumstats.MACfilt.txt.gz |
|  | 31427789 | 2019 | Stomach or abdominal pain | UKB2 (EUR) | 385698 | 0.0179 | GWASA_ID_3573-Stomach_or_Abdominal_Pain | https://atlas.ctglab.nl/ukb2_sumstats/6159_5_logistic.EUR.sumstats.MACfilt.txt.gz |
|  | 24097068 | 2013 | Total cholesterol | EUR | 188577 | 0.1508 | GWASA_ID_73-Total_cholesterol | http://csg.sph.umich.edu/abecasis/public/lipids2013/jointGwasMc_TC.txt.gz |
|  | 22982992 | 2012 | Variability of Height | EUR | 133154 | 0.0108 | GWASA_ID_116-Variability_of_Height | http://portals.broadinstitute.org/collaboration/giant/images/b/b0/GIANT_Yang2012Nature_publicrelease_HapMapCeuFreq_Height.txt.gz |
|  | 31427789 | 2019 | Waist circumference | UKB2 (EUR) | 385932 | 0.1803 | GWASA_ID_3185-Waist_circumference | https://atlas.ctglab.nl/ukb2_sumstats/f.48.0.0_res.EUR.sumstats.MACfilt.txt.gz |
|  | 30239722 | 2018 | Waist-hip ratio | UKB2 (EUR meta) | 697734 | 0.1246 | GWASA_ID_4077-Waist_hip_ratio | https://zenodo.org/record/1251813/files/whr.giant-ukbb.meta-analysis.combined.23May2018.txt.gz?download=1 |
| Personality | 31427789 | 2019 | Anxious | UKB2 (EUR) | 376411 | 0.0736 | GWASA_ID_3290-Anxious | https://atlas.ctglab.nl/ukb2_sumstats/f.1980.0.0_logistic.EUR.sumstats.MACfilt.txt.gz |
|  | 29942085 | 2018 | Depressive affect | UKB2 (EUR) | 357957 | 0.088 | GWASA_ID_3797-Depressive_Affect | https://ctg.cncr.nl/documents/p1651/sumstats_depressed_affect_ctg_format.txt.gz |
|  | 26362575 | 2016 | Extraversion | EUR | 63661 | 0.0497 | GWASA_ID_33-Extraversion_IRT | https://www.dropbox.com/s/bk2jn41vrfl3zna/GPC-2.EXTRAVERSION.zip?dl=0 |
|  | 29500382 | 2018 | Fed-up | UKB2 (EUR) | 266208 | 0.0647 | GWASA_ID_3995-Fed_Up | https://ctg.cncr.nl/documents/p1651/sumstats_neuro_f1960_ctg_format.txt.gz |
|  | 29500382 | 2018 | Guilty feelings | UKB2 (EUR) | 265139 | 0.049 | GWASA_ID_4002-Guilty_Feelings | https://ctg.cncr.nl/documents/p1651/sumstats_neuro_f2030_ctg_format.txt.gz |
|  | 31427789 | 2019 | Happiness | UKB2 (EUR) | 128677 | 0.0622 | GWASA_ID_3385-Happiness | https://atlas.ctglab.nl/ukb2_sumstats/f.4526.0.0_res.EUR.sumstats.MACfilt.txt.gz |
|  | 31427789 | 2019 | Irritability | UKB2 (EUR) | 369232 | 0.063 | GWASA_ID_3286-Irritability | https://atlas.ctglab.nl/ukb2_sumstats/f.1940.0.0_logistic.EUR.sumstats.MACfilt.txt.gz |
|  | 29500382 | 2018 | Loneliness | UKB2 (EUR) | 267190 | 0.0351 | GWASA_ID_4001-Loneliness | https://ctg.cncr.nl/documents/p1651/sumstats_neuro_f2020_ctg_format.txt.gz |
|  | 29500382 | 2018 | Miserableness | UKB2 (EUR) | 267050 | 0.0587 | GWASA_ID_3992-Miserableness | https://ctg.cncr.nl/documents/p1651/sumstats_neuro_f1930_ctg_format.txt.gz |
|  | 29500382 | 2018 | Mood swings | UKB2 (EUR) | 265382 | 0.067 | GWASA_ID_3991-Mood_Swings | https://ctg.cncr.nl/documents/p1651/sumstats_neuro_f1920_ctg_format.txt.gz |
|  | 31427789 | 2019 | Nervous | UKB2 (EUR) | 376368 | 0.0623 | GWASA_ID_3289-Nervous | https://atlas.ctglab.nl/ukb2_sumstats/f.1970.0.0_logistic.EUR.sumstats.MACfilt.txt.gz |
|  | 31427789 | 2019 | Neuroticism | UKB2 (EUR) | 312740 | 0.1086 | GWASA_ID_3417-Neuroticism | https://atlas.ctglab.nl/ukb2_sumstats/f.20127.0.0_res.EUR.sumstats.MACfilt.txt.gz |
|  | 31427789 | 2019 | Risk taking | UKB2 (EUR) | 372651 | 0.0534 | GWASA_ID_3296-Risk_Taking | https://atlas.ctglab.nl/ukb2_sumstats/f.2040.0.0_logistic.EUR.sumstats.MACfilt.txt.gz |
|  | 29500382 | 2018 | Sensitivity | UKB2 (EUR) | 264144 | 0.0594 | GWASA_ID_3994-Sensitivity | https://ctg.cncr.nl/documents/p1651/sumstats_neuro_f1950_ctg_format.txt.gz |
|  | 27089181 | 2016 | Subjective well being | EUR | 298420 | 0.0251 | GWASA_ID_54-Subjective_Well_Being | http://ssgac.org/documents/SWB_Full.txt.gz |
|  | 29500382 | 2018 | Suffer from nerves | UKB2 (EUR) | 262321 | 0.0417 | GWASA_ID_4000-Suffer_from_Nerves | https://ctg.cncr.nl/documents/p1651/sumstats_neuro_f2010_ctg_format.txt.gz |
|  | 31427789 | 2019 | Tense | UKB2 (EUR) | 374129 | 0.0536 | GWASA_ID_3291-Tense | https://atlas.ctglab.nl/ukb2_sumstats/f.1990.0.0_logistic.EUR.sumstats.MACfilt.txt.gz |
|  | 29942085 | 2018 | Worry | UKB2 (EUR) | 348219 | 0.0894 | GWASA_ID_3798-Worry_Subcluster | https://ctg.cncr.nl/documents/p1651/sumstats_worry_ctg_format.txt.gz |
|  | 31427789 | 2019 | Worry too long after embarrassment | UKB2 (EUR) | 370660 | 0.0603 | GWASA_ID_3292-Worry_too_Long_After_Embarassment | https://atlas.ctglab.nl/ukb2_sumstats/f.2000.0.0_logistic.EUR.sumstats.MACfilt.txt.gz |
| Psychiatric Disorders | 30478444 | 2019 | Attention deficit hyperactivity disorder | EUR | 53293 | 0.2359 | GWASA_ID_2-Attention_deficit_hyperactivity_disorder | https://www.med.unc.edu/pgc/results-and-downloads |
|  | 29906448 | 2018 | Bipolar disorder | EUR | 74194 | 0.1991 | GWASA_ID_4039-Bipolar_disorder | https://www.med.unc.edu/pgc/results-and-downloads/downloads |
|  | 31427789 | 2019 | Bipolar/Major depression | UKB2 (EUR) | 93296 | 0.0623 | GWASA_ID_3416-Bipolar_and_major_depression_status | https://atlas.ctglab.nl/ukb2_sumstats/f.20126.0.0_logistic.EUR.sumstats.MACfilt.txt.gz |
|  | 29662059 | 2018 | Depression | UKB2 (EUR) | 322580 | 0.0622 | GWASA_ID_4011-Depression | https://datashare.is.ed.ac.uk/bitstream/handle/10283/3036/UKBiobank_broad_12Jan18.txt?sequence=1&isAllowed=y |
|  | 27089181 | 2016 | Depressive symptoms | EUR | 161460 | 0.0475 | GWASA_ID_56-Depressive_Symptoms | http://ssgac.org/documents/DS_Full.txt.gz |
|  | 29700475 | 2018 | Major depressive disorder | UKB1 (EUR meta) | 173005 | 0.0774 | GWASA_ID_4014-Major_Depressive_Disorder | https://www.med.unc.edu/pgc/results-and-downloads/downloads |
|  | 28761083 | 2018 | Obsessive compulsive disorder | EUR+AFR | 9725 | 0.3237 | GWASA_ID_4042-Obsessive_compulsive_disorder | https://www.med.unc.edu/pgc/results-and-downloads/downloads |
|  | 28439101 | 2017 | Posttraumatic stress disorder | EUR+AFR+others | 19884 | 0.0402 | GWASA_ID_16-Posttraumatic_stress_disorder | https://www.med.unc.edu/pgc/results-and-downloads |
|  | 29906448 | 2018 | Schizophrenia | EUR | 87491 | 0.3199 | GWASA_ID_4038-Schizophrenia | https://www.med.unc.edu/pgc/results-and-downloads/downloads |
| Sleep | 31427789 | 2019 | Chronotype | UKB2 (EUR) | 345148 | 0.1119 | GWASA_ID_3230-Chronotype | https://atlas.ctglab.nl/ukb2_sumstats/f.1180.0.0_res.EUR.sumstats.MACfilt.txt.gz |
|  | 31427789 | 2019 | Insomnia | UKB2 (EUR) | 386078 | 0.0605 | GWASA_ID_3232-Insomnia | https://atlas.ctglab.nl/ukb2_sumstats/f.1200.0.0_res.EUR.sumstats.MACfilt.txt.gz |
| Substance | 30643258 | 2019 | Alcohol intake | UKB2 (EUR) | 414343 | 0.0687 | GWASA_ID_4069-Drinks_Per_Week | https://www.dropbox.com/s/7hjxdhlxlwa482n/DRINKS_PER_WEEK_GWAS.txt?dl=1 |
|  | 31427789 | 2019 | Alcohol intake frequency | UKB2 (EUR) | 386082 | 0.0717 | GWASA_ID_3261-Alcohol_Intake_Frequency | https://atlas.ctglab.nl/ukb2_sumstats/f.1558.0.0_res.EUR.sumstats.MACfilt.txt.gz |
|  | 31427789 | 2019 | Ever taken cannabis | UKB2 (EUR) | 126632 | 0.0646 | GWASA_ID_3744-Cannabis_use | https://atlas.ctglab.nl/ukb2_sumstats/f.20453.0.0_res.EUR.sumstats.MACfilt.txt.gz |
|  | 31427789 | 2019 | Ever vs never drinkers | UKB2 (EUR) | 386082 | 0.0156 | GWASA_ID_3656-Ever_vs_Never_Drinkers | https://atlas.ctglab.nl/ukb2_sumstats/20117_0_logistic.EUR.sumstats.MACfilt.txt.gz |
|  | 30643258 | 2019 | Ever vs never smokers | UKB2 (EUR meta) | 518633 | 0.0877 | GWASA_ID_4070-Ever_vs_Never_Smokers | https://www.dropbox.com/s/o7wgwhnhjgt3eyn/EVER_SMOKER_GWAS_MA_UKB%2BTAG.txt?dl=1 |
|  | 31427789 | 2019 | Former vs current drinkers | UKB2 (EUR) | 373560 | 0.0113 | GWASA_ID_3657-Former_vs_Current_Drinkers | https://atlas.ctglab.nl/ukb2_sumstats/20117_1_2_logistic.EUR.sumstats.MACfilt.txt.gz |
| Temperament | 31427789 | 2019 | Frequency of depressed mood in last 2 weeks | UKB2 (EUR) | 370017 | 0.0387 | GWASA_ID_3297-Frequency_of_Depressed_Mood | https://atlas.ctglab.nl/ukb2_sumstats/f.2050.0.0_res.EUR.sumstats.MACfilt.txt.gz |
|  | 31427789 | 2019 | Frequency of tenseness / restlessness in last 2 weeks | UKB2 (EUR) | 371869 | 0.0405 | GWASA_ID_3299-Frequency_of_Tenseness | https://atlas.ctglab.nl/ukb2_sumstats/f.2070.0.0_res.EUR.sumstats.MACfilt.txt.gz |
|  | 31427789 | 2019 | Frequency of tiredness / lethargy in last 2 weeks | UKB2 (EUR) | 375053 | 0.0518 | GWASA_ID_3300-Frequency_of_Tiredness | https://atlas.ctglab.nl/ukb2_sumstats/f.2080.0.0_res.EUR.sumstats.MACfilt.txt.gz |
|  | 31427789 | 2019 | Frequency of unenthusiasm / disinterest in last 2 weeks | UKB2 (EUR) | 373833 | 0.0329 | GWASA_ID_3298-Frequency_of_Unenthusiasm | https://atlas.ctglab.nl/ukb2_sumstats/f.2060.0.0_res.EUR.sumstats.MACfilt.txt.gz |
|  | 31427789 | 2019 | Seen a psychiatrist for nerves, anxiety, tension or depression | UKB2 (EUR) | 384700 | 0.0301 | GWASA_ID_3302-Seen_a_psychiatrist_for_nerves_anxiety_tension_or_depression | https://atlas.ctglab.nl/ukb2_sumstats/f.2100.0.0_logistic.EUR.sumstats.MACfilt.txt.gz |
|  | 31427789 | 2019 | Seen doctor (GP) for nerves, anxiety, tension or depression | UKB2 (EUR) | 383771 | 0.0589 | GWASA_ID_3301-Seen_doctor_GP_for_nerves_anxiety_tension_or_depression | https://atlas.ctglab.nl/ukb2_sumstats/f.2090.0.0_logistic.EUR.sumstats.MACfilt.txt.gz |

**eTable 3. Validation results for 44 PGS with significant associations to MDD identified in the discovery set.**

| **Category** | **PMID** | **uniqTrait** | **Filename** | **trMDD vs non-MDD** | | | | **TRD vs non-MDD** | | | | **TRD vs trMDD** | |
| --- | --- | --- | --- | --- | --- | --- | --- | --- | --- | --- | --- | --- | --- |
|  |  |  |  | *European set* | | *diverse set* | | *European set* | | *diverse set* | | *all population* | |
|  |  |  |  | beta | p-value | beta | p-value | beta | p-value | beta | p-value | beta | p-value |
| Education and Cognition | 30038396 | Educational attainment | GWASA_ID_4066__Educational_Attainment | -9.61E-02 | 1.02E-27 | -8.05E-02 | 5.44E-07 | -2.24E-01 | 2.93E-41 | -1.16E-01 | 4.45E-04 | -1.31E-01 | 3.87E-12 |
|  | 30038396 | Intelligence | GWASA_ID_4067__Cognitive_Performance | -4.34E-02 | 2.57E-07 | -2.49E-02 | 4.27E-02 | -1.35E-01 | 1.61E-17 | -4.87E-02 | 5.38E-02 | -8.21E-02 | 2.10E-07 |
| Metabolic, Somatic Complaints, and Inflammations | 31427789 | Back pain | GWASA_ID_3572__Back_Pain | 6.49E-02 | 1.19E-09 | 7.21E-02 | 2.98E-03 | 9.43E-02 | 2.80E-06 | 5.58E-02 | 2.58E-01 | 3.08E-02 | 3.49E-01 |
|  | 30239722 | Body Mass Index | GWASA_ID_4074__Body_Mass_Index | 1.22E-01 | 2.19E-47 | 1.67E-01 | 1.76E-11 | 1.49E-01 | 7.23E-21 | 1.45E-01 | 4.48E-03 | 3.19E-02 | 2.05E-01 |
|  | 6218410 | CRP | HapMap_GWAS_CRP_AJHG | 3.57E-02 | 2.79E-05 | 3.09E-03 | 8.31E-01 | 2.31E-02 | 1.50E-01 | 3.52E-02 | 2.35E-01 | -2.52E-03 | 8.79E-01 |
|  | 31427789 | Headache | GWASA_ID_3570__Headache | 7.03E-02 | 1.18E-08 | 8.06E-02 | 1.31E-02 | 1.52E-01 | 7.08E-11 | 7.71E-02 | 2.46E-01 | 1.00E-01 | 1.09E-02 |
|  | 31427789 | Hip pain | GWASA_ID_3574__Hip_Pain | 8.23E-02 | 8.23E-16 | 7.81E-02 | 6.23E-06 | 1.34E-01 | 4.04E-12 | 7.20E-02 | 4.13E-02 | 5.02E-02 | 2.32E-02 |
|  | 31427789 | Knee pain | GWASA_ID_3575__Knee_Pain | 7.77E-02 | 9.20E-18 | 3.40E-02 | 3.93E-03 | 1.38E-01 | 8.85E-16 | 4.48E-02 | 6.30E-02 | 4.88E-02 | 1.21E-03 |
|  | 31427789 | Neck or shoulder pain | GWASA_ID_3571__Neck_or_Shoulder_Pain | 9.45E-02 | 1.18E-17 | 1.03E-01 | 2.80E-05 | 1.46E-01 | 2.89E-12 | 7.30E-02 | 1.45E-01 | 5.83E-02 | 1.05E-01 |
|  | 31427789 | Stomach or abdominal pain | GWASA_ID_3573__Stomach_or_Abdominal_Pain | 6.40E-02 | 2.07E-09 | 1.18E-01 | 6.79E-08 | 1.52E-01 | 4.13E-14 | 6.43E-02 | 1.47E-01 | 8.64E-02 | 4.24E-03 |
|  | 31427789 | Waist circumference | GWASA_ID_3185__Waist_circumference | 1.62E-01 | 1.10E-42 | 1.35E-01 | 8.19E-12 | 1.93E-01 | 5.78E-18 | 1.10E-01 | 6.51E-03 | 3.12E-02 | 2.42E-01 |
|  | 30239722 | Waist-hip ratio | GWASA_ID_4077__Waist_hip_ratio | 9.88E-02 | 7.57E-31 | 6.50E-02 | 1.08E-06 | 1.37E-01 | 1.72E-17 | 5.29E-02 | 5.25E-02 | 3.18E-02 | 6.99E-02 |
| Personality | 31427789 | Anxious | GWASA_ID_3290__Anxious | 3.60E-02 | 1.42E-04 | 4.70E-02 | 3.23E-03 | 5.39E-02 | 2.53E-03 | 9.52E-03 | 7.70E-01 | 4.10E-03 | 8.39E-01 |
|  | 29942085 | Depressive affect | GWASA_ID_3797__Depressive_Affect | 1.48E-01 | 4.81E-69 | 1.31E-01 | 1.23E-17 | 2.45E-01 | 1.27E-53 | 1.67E-01 | 8.96E-08 | 8.46E-02 | 3.91E-08 |
|  | 29500382 | Fed-up | GWASA_ID_3995__Fed_Up | 1.29E-01 | 1.36E-52 | 9.08E-02 | 1.45E-09 | 2.20E-01 | 9.03E-44 | 1.48E-01 | 1.51E-06 | 8.46E-02 | 3.30E-08 |
|  | 29500382 | Guilty feelings | GWASA_ID_4002__Guilty_Feelings | 8.71E-02 | 4.44E-25 | 6.44E-02 | 6.93E-07 | 1.40E-01 | 1.03E-18 | 1.07E-01 | 5.50E-05 | 5.45E-02 | 6.71E-04 |
|  | 31427789 | Happiness | GWASA_ID_3385__Happiness | -6.88E-02 | 1.46E-07 | -6.42E-02 | 2.33E-02 | -9.76E-02 | 7.97E-05 | -1.04E-01 | 7.20E-02 | -3.57E-02 | 3.50E-01 |
|  | 31427789 | Irritability | GWASA_ID_3286__Irritability | 7.18E-02 | 6.36E-09 | 6.04E-02 | 4.45E-03 | 1.39E-01 | 2.62E-09 | 8.02E-02 | 6.53E-02 | 5.66E-02 | 3.41E-02 |
|  | 29500382 | Loneliness | GWASA_ID_4001__Loneliness | 9.45E-02 | 3.14E-29 | 8.36E-02 | 1.19E-06 | 1.38E-01 | 3.41E-18 | 1.79E-01 | 3.99E-07 | 6.46E-02 | 4.28E-04 |
|  | 29500382 | Miserableness | GWASA_ID_3992__Miserableness | 7.33E-02 | 3.91E-18 | 6.42E-02 | 5.19E-08 | 1.20E-01 | 3.59E-14 | 6.09E-02 | 1.15E-02 | 3.50E-02 | 2.14E-02 |
|  | 29500382 | Mood swings | GWASA_ID_3991__Mood_Swings | 1.35E-01 | 6.47E-58 | 1.25E-01 | 1.99E-15 | 2.25E-01 | 1.50E-45 | 1.78E-01 | 3.61E-08 | 8.24E-02 | 1.12E-07 |
|  | 31427789 | Nervous | GWASA_ID_3289__Nervous | 6.25E-02 | 4.85E-12 | 3.52E-02 | 1.55E-02 | 1.02E-01 | 2.20E-09 | 3.55E-02 | 2.34E-01 | 2.83E-02 | 9.11E-02 |
|  | 31427789 | Neuroticism | GWASA_ID_3417__Neuroticism | 1.51E-01 | 1.04E-53 | 1.48E-01 | 2.50E-13 | 2.54E-01 | 4.06E-43 | 2.15E-01 | 2.00E-07 | 1.12E-01 | 2.66E-07 |
|  | 29500382 | Sensitivity | GWASA_ID_3994__Sensitivity | 1.00E-01 | 1.50E-32 | 1.06E-01 | 1.01E-14 | 1.73E-01 | 1.89E-27 | 1.25E-01 | 7.69E-06 | 5.97E-02 | 6.10E-05 |
|  | 27089181 | Subjective well being | GWASA_ID_54__Subjective_Well_Being | -6.12E-02 | 6.85E-13 | -4.50E-02 | 1.27E-03 | -9.90E-02 | 7.15E-10 | -8.36E-02 | 3.51E-03 | -3.75E-02 | 1.53E-02 |
|  | 29500382 | Suffer from nerves | GWASA_ID_4000__Suffer_from_Nerves | 7.45E-02 | 7.67E-19 | 3.55E-02 | 2.47E-03 | 1.40E-01 | 1.04E-18 | 3.70E-02 | 1.23E-01 | 4.79E-02 | 9.53E-04 |
|  | 31427789 | Tense | GWASA_ID_3291__Tense | 1.22E-01 | 1.37E-15 | 1.43E-01 | 2.19E-04 | 2.10E-01 | 3.25E-13 | 2.03E-01 | 1.03E-02 | 1.24E-01 | 1.56E-02 |
|  | 29942085 | Worry | GWASA_ID_3798__Worry_Subcluster | 6.65E-02 | 2.70E-15 | 5.17E-02 | 9.14E-06 | 1.19E-01 | 6.28E-14 | 5.13E-02 | 3.16E-02 | 3.71E-02 | 1.05E-02 |
|  | 31427789 | Worry too long after embarrassment | GWASA_ID_3292__Worry_too_Long_After_Embarassment | 7.09E-02 | 7.52E-13 | 7.75E-02 | 4.40E-04 | 9.42E-02 | 4.26E-07 | 9.06E-02 | 4.51E-02 | 2.30E-02 | 3.32E-01 |
| Psychiatric Disorders | 30478444 | Attention deficit hyperactivity disorder | GWASA_ID_2__Attention_deficit_hyperactivity_disorder | 6.83E-02 | 7.35E-16 | 5.07E-02 | 2.46E-03 | 1.11E-01 | 3.88E-12 | 4.14E-02 | 2.26E-01 | 4.69E-02 | 3.37E-02 |
|  | 31427789 | Bipolar/Major depression | GWASA_ID_3416__Bipolar_and_major_depression_status | 8.70E-02 | 8.12E-21 | 7.60E-02 | 1.30E-05 | 7.83E-02 | 7.65E-06 | 6.61E-02 | 6.35E-02 | -1.11E-02 | 5.74E-01 |
|  | 29662059 | Depression | GWASA_ID_4011__Depression | 7.51E-02 | 3.76E-19 | 6.92E-02 | 1.02E-07 | 8.72E-02 | 3.50E-08 | 2.85E-02 | 2.83E-01 | -2.52E-03 | 8.74E-01 |
|  | 27089181 | Depressive symptoms | GWASA_ID_56__Depressive_Symptoms | 1.03E-01 | 5.76E-34 | 6.43E-02 | 1.90E-06 | 1.57E-01 | 8.41E-23 | 8.25E-02 | 2.81E-03 | 5.01E-02 | 2.11E-03 |
|  | 29700475 | Major depressive disorder | GWASA_ID_4014__Major_Depressive_Disorder | 8.58E-02 | 2.27E-24 | 3.45E-02 | 2.91E-02 | 9.63E-02 | 1.31E-09 | 7.39E-02 | 2.11E-02 | 2.21E-02 | 2.12E-01 |
| Sleep | 31427789 | Insomnia | GWASA_ID_3232__Insomnia | 7.63E-02 | 4.40E-17 | 7.13E-02 | 3.65E-05 | 1.78E-01 | 3.21E-25 | 1.26E-01 | 3.81E-04 | 1.03E-01 | 5.57E-08 |
| Substance | 31427789 | Alcohol intake frequency | GWASA_ID_3261__Alcohol_Intake_Frequency | -5.80E-02 | 9.49E-06 | -8.97E-02 | 4.52E-03 | -5.96E-02 | 1.62E-02 | -9.27E-02 | 1.50E-01 | 7.29E-03 | 8.58E-01 |
|  | 30643258 | Ever vs never smokers | GWASA_ID_4070__Ever_vs_Never_Smokers | 7.12E-02 | 2.68E-17 | 3.47E-02 | 2.87E-03 | 8.79E-02 | 2.88E-08 | 2.82E-02 | 2.36E-01 | 1.26E-02 | 3.80E-01 |
|  | 31427789 | Former vs current drinkers | GWASA_ID_3657__Former_vs_Current_Drinkers | 9.18E-02 | 1.11E-14 | 5.11E-02 | 6.15E-03 | 1.17E-01 | 2.04E-07 | 5.22E-02 | 1.71E-01 | 3.16E-02 | 2.21E-01 |
| Temperament | 31427789 | Frequency of depressed mood in last 2 weeks | GWASA_ID_3297__Frequency_of_Depressed_Mood | 1.48E-01 | 1.08E-46 | 2.03E-01 | 1.71E-10 | 2.40E-01 | 5.91E-35 | 3.13E-01 | 1.63E-06 | 1.89E-01 | 4.46E-06 |
|  | 31427789 | Frequency of tenseness / restlessness in last 2 weeks | GWASA_ID_3299__Frequency_of_Tenseness | 1.42E-01 | 1.40E-29 | 2.54E-01 | 5.14E-11 | 2.84E-01 | 5.10E-33 | 3.58E-01 | 6.18E-06 | 2.49E-01 | 2.53E-07 |
|  | 31427789 | Frequency of tiredness / lethargy in last 2 weeks | GWASA_ID_3300__Frequency_of_Tiredness | 1.34E-01 | 5.57E-47 | 1.33E-01 | 7.16E-12 | 2.62E-01 | 1.62E-50 | 1.52E-01 | 1.34E-04 | 1.42E-01 | 1.31E-09 |
|  | 31427789 | Frequency of unenthusiasm / disinterest in last 2 weeks | GWASA_ID_3298__Frequency_of_Unenthusiasm | 1.15E-01 | 1.36E-39 | 1.18E-01 | 3.12E-06 | 2.22E-01 | 4.42E-41 | 2.42E-01 | 2.58E-06 | 2.04E-01 | 4.26E-10 |
|  | 31427789 | Seen a psychiatrist for nerves, anxiety, tension or depression | GWASA_ID_3302__Seen_a_psychiatrist_for_nerves_anxiety_tension_or_depression | 1.22E-01 | 3.58E-22 | 1.85E-01 | 2.94E-09 | 1.52E-01 | 1.60E-10 | 2.03E-01 | 1.42E-03 | 3.79E-02 | 3.32E-01 |
|  | 31427789 | Seen doctor (GP) for nerves, anxiety, tension or depression | GWASA_ID_3301__Seen_doctor_GP_for_nerves_anxiety_tension_or_depression | 1.57E-01 | 6.64E-64 | 1.61E-01 | 3.95E-16 | 2.08E-01 | 1.53E-32 | 2.01E-01 | 6.94E-07 | 5.84E-02 | 5.95E-03 |
